# School-based caries prevention using silver diamine fluoride: A pragmatic randomized trial in low-income rural children

**DOI:** 10.1101/2024.06.05.24308499

**Authors:** Ryan Richard Ruff, Richard Niederman

## Abstract

**Background:** Dental caries is the world’s most prevalent noncommunicable disease, disproportionately affecting children from low-income families and rural geographic areas.

**Methods:** The CariedAway 3.0 study was a cluster-randomized pragmatic non-inferiority trial comparing silver diamine fluoride (SDF) to sealants and atraumatic restorations (ART) for the prevention and control of dental caries. All participants also received fluoride varnish. Analysis consisted of mixed-effects logistic regression for caries prevalence and weighted least squares and mixed-effects negative binomial regression for caries incidence. A non-inferiority margin of 10% for the difference between groups was used. Dental caries was defined as an ICDAS score of four or greater.

**Results:** A total of 3345 children were enrolled across 35 schools; however there was a large proportion of children who were noncompliant and received external dental care over the course of the trial. In adjusted analyses of compliant participants (n=1083; 543 in the SDF group and 540 in the sealant and ART group), there was no difference in the weighted risk difference between treatment groups (B=0.003, 95% CI = -0.0001, 0.0008). The odds of caries prevalence was elevated in the SDF group in longitudinal analyses (OR = 1.35, 95% CI = 0.86, 2.11) but was not significant and was below the non-inferiority margin. There were no significant differences between groups for caries incidence in adjusted models (IRR = 1.19, 95% CI = 0.81, 1.74). Results for intent to treat analyses were similar to that of per-protocol.

**Discussion:** In this school-based clinical trial, the prevalence of dental caries in children treated with SDF and fluoride varnish was non-inferior compared to those treated with sealants, ART, and fluoride varnish, although the overall risk was slightly higher. Unfortunately, a high rate of dropout and participant noncompliance was observed, likely due to the impacts of COVID-19 on study procedures. As a result, observed effects may be unreliable beyond the short-term.

**Trial Registration:** NCT03448107

## Introduction

Dental caries is a public health crisis and is the world’s most common non-communicable disease ^1^, with more than 2.5 billion people having caries in the primary or permanent dentition ^2^. In addition to infection and pain, poor oral health is associated with reduced academic performance ^3^ and lower quality of life ^4^. Over fifty years ago, the “inverse care law” demonstrated that as the need for healthcare increases in a population, access decreases ^5^. This inequity largely persists today, with those living in rural areas facing lower access to dental care ^6^ and being less likely to have dental insurance ^7,8^. As a result, these populations are disproportionately affected by caries and other oral diseases ^9,10^. In recognition of these needs, the World Health Organization’s Global Oral Health Action Plan includes calls for evidence-based interventions to prevent oral diseases, innovative workforce models for oral health, and increased access to essential curative oral health care ^11^.

Common interventions to reduce oral health access barriers in rural populations include mobile dental clinics, school-based caries prevention, teledentistry, and dental outreach programs and educational initiatives ^6^. School-based caries prevention can successfully promote healthy behaviors and reduce dental caries by providing screening and oral health education ^12^, fluoride supplementation and outpatient services ^13^, dental sealants ^14^, and atraumatic restorations ^15^, forming part of the Whole School, Whole Community, Whole Child model ^16^ through the development of interventions in collaboration with the educational sector to leverage schools in advancing oral health ^17^. However, the implementation and sustainability of school caries prevention remains a consistent challenge ^18^.

In 2022, the WHO added silver diamine fluoride (SDF) to its Model List of Essential Medicines ^19^. As an effective strategy to control and manage existing dental caries ^20^, SDF is cost-efficient ^21^, fast to apply ^22^, and can be effectively administered by both dental hygienists and nurses ^23^. More recently, data indicates that SDF is also effective in preventing dental caries ^24,25^. Introducing silver diamine fluoride into school caries prevention may dramatically increase access to care and obviate existing implementation barriers. The objective of the *CariedAway 3.0* trial was to assess the feasibility and effectiveness of integrating silver diamine fluoride into a rural, school-based caries prevention program ^26^. We present findings on the prevalence and incidence of dental caries in predominantly low-income children receiving SDF, compared to dental sealants.

## Methods

### Design and Participants

The CariedAway 3.0 (“CariedAway”) study was a longitudinal, cluster-randomized, pragmatic non-inferiority trial conducted from 20 September 2017 to 30 June 2023. The study is reported using the Enhancing the Quality and Transparency of Health Research (EQUATOR) guidelines, was registered at www.clinicaltrials.gov (#NCT03448107), and was approved by the New York University School of Medicine Institutional Review Board (#i17-01221). A trial protocol is available ^26^.

CariedAway was conducted in eligible primary schools in five counties in New Hampshire, USA, with a student population consisting of predominantly rural white children. Any primary school with a previously employed caries prevention program, with at least 40% of enrolled children being from low-income families (“Title 1”), and located in a health professional shortage area was eligible for inclusion. Within participating schools, all children with informed consent and assent were eligible to receive care, however inclusion into the research study was restricted to children between the ages of 5 and 12 years.

### Clinical Assessment

All study participants received a dental screening, toothbrush cleaning, and oral hygiene instruction, followed by the allocated interventions. Screening and treatment were provided in a room designated by the school (e.g., library, empty classroom, or conference room). Visual-tactile assessments of all tooth surfaces were performed with the child supine in a portable dental chair using a disposable mirror, disposable plastic explorer, and dental light. No hard or soft tissue was removed from carious lesions beyond cleaning with a toothbrush or explorer.

### Interventions

This two-arm trial included an experimental condition of silver diamine fluoride and an active comparator of dental sealants and atraumatic restorations. Both groups also received fluoride varnish. The original protocol stipulated a twice-yearly dosage frequency.

For the experimental treatment, one drop (0.05 ml) of silver diamine fluoride (Elevate Oral Care, Advantage Arrest Silver Diamine Fluoride 38%, 2.24 F-ion mg/dose) was dispensed for each participant. Posterior tooth surfaces were dried, after which SDF was applied using a micro-brush to all asymptomatic carious lesions and to all pits and fissures on primary and permanent posterior teeth for thirty seconds. Fluoride varnish (Benco Iris 5% NaF, or Elevate Oral Care FluoriMax 2.5% NaF for those with nut allergies) was then applied to all teeth to mask the bitter, metallic aftertaste of SDF.

For the active control, pits and fissures on all posterior teeth (primary and permanent) were treated for 10 seconds with 20% polyacrylic acid (GC Cavity Conditioner), dried with a cotton roll or 2”x2” swab. Glass ionomer capsules (GC Equia Forte), triturated for 8-10 seconds, was then delivered with a capsule applier (CG Capsule Applier). A moistened finger then pressed the glass ionomer into the pits and fissures, followed by a moistened, cotton-tipped applicator to ensure margin sealing. Subsequent flossing removed any excess interproximal material. All asymptomatic, frank cavitated lesions were treated similarly and simultaneously to the pits and fissures, with the same material and technique. For Class II lesions, the applicator tip was placed at the base of the interproximal lesion, moved occlusally, and then mesially or distally. Moist finger pressure and moist cotton tip applicator pressure secured the glass ionomer in a manner similar to sealants. Subsequent interproximal flossing removed excess interproximal material. Fluoride varnish (Benco Iris 5% NaF, or Elevate Oral Care FluoriMax 2.5% NaF for those with nut allergies) was then placed on all teeth.

### Diagnosis and Outcomes

The primary outcome for the CariedAway trial was dental caries, assessed via the total number of participants presenting with new caries and the number of sound teeth treated with either SDF or sealants and ART that developed new caries over time. At each study observation, each tooth surface was assessed as sound (no evidence of decay), sealed (fossa, pits, and fissures visibly and completely sealed), restored (including fillings with amalgam, resin, glass ionomer, or crowns), decayed, or arrested decay (hard black surface). Decay diagnosis was determined by frank cavitated lesions defined as a score of 4 or higher on the International Caries Detection and Assessment System ^27^ (ICDAS).

### Randomization and Masking

Schools participating in CariedAway were block-randomized to either condition using a random number generator. As a pragmatic trial, blinding was not feasible.

### Impact of COVID-19

Due to COVID-19, schools participating in the CariedAway trial were closed from 15 March 2020 to 1 January 2021 and all study procedures were suspended. As a result, the biannual treatment schedule was unable to be maintained. Additionally, study enrollment and follow-up with participants was affected, as schools and subjects were either unable to be enrolled, needed to re-enroll, or were otherwise lost to follow-up due to the extended duration between study observations.

### Statistical Analysis

Participants were first ordered sequentially by observational period and descriptive statistics for sociodemographic (e.g., sex, race/ethnicity, age) and clinical (baseline decay, preexisting sealants) variables were computed (N/% for categorical indicators, mean/SD for continuous indicators). For teeth treated with either SDF or sealants and ART, we first calculated the person-level prevalence of untreated decay at each observation and the total number of decayed teeth diagnosed, estimating the crude incidence rate. Given the variable rates of follow-up in study participants, we computed the rate difference using weighted least squares regression, adjusting for age and sex, and report estimates of any new decay that was observed ^28^. As decay could have occurred at any point between observations, we subsequently modeled caries prevalence and the number of new caries by study observation using multilevel mixed-effects logistic and negative binomial regression, respectively, including random intercepts for school and patient. Longitudinal analyses further adjusted for the presence of baseline decay and the number of teeth with verified fillings, reflecting treatment for caries that was received outside of the study. Since the prevalence of subjects with caries is a negative outcome, we compared the upper limit of the (1-2α) confidence interval of silver diamine fluoride to the inverse non-inferiority margin on the odds ratio scale (2.22) ^29^.

We also conducted a series of sensitivity analyses. We first compared the prevalence of caries for all participants six months after their baseline treatment, which avoids disparities in attrition between groups that was observed in later visits. Differences in caries prevalence used two-sample proportion tests with bootstrapped confidence intervals (10,000 replications), adjusting for the clustering effect of schools. In multilevel analyses, we then considered multiple interaction effects including treatment group and baseline decay, observational period and baseline decay, and the quadratic effect of the observational period. Finally, although participant crossover in this study was minimal there was a substantial loss to follow-up and treatment noncompliance, the latter of which being due to receiving dental care outside of the school-based program. To supplement estimates for the potential causal effect of treatment, we compared study dropouts between treatment groups and conducted an intent to treat analysis ^30^. For this latter approach, we first estimated inverse probability weights using regression conditioning on continuation, followed by weighted generalized estimating equations for caries prevalence. Analysis was performed in Stata v18 (StataCorp, LLC) and R v 4.3 (R Foundation).

## Results

A total of 35 schools were enrolled and randomized. Within these schools, 3670 children were enrolled of which 3345 met inclusion criteria for evaluation. There were 1558 (46.6%) participants in the SDF group and 1787 (53.4%) in the sealant and ART group (Table 1, Figure 1). The intraclass correlation for school-level clustering of decay prevalence was 0.019. The overall baseline prevalence of untreated caries in treatable teeth was 14.9%, or 14.1% (95% CI = 12.5, 15.9) in the SDF group and 15.5% (95% CI = 13.8, 17.1) in the sealant and ART group. Over 70% of the sample did not report race/ethnicity, and 26% of participants were white. The average age at baseline was 7.2y (SD=1.8). The prevalence of preexisting sealants was 49.8% (44.0% in the SDF group, 54.9% in the sealant and ART group). These subjects were removed prior to adjusted analyses (n=1665).

**Table 1:** Participant descriptive statistics.

|  | Overall |  | SDF |  | Sealants |  |
| --- | --- | --- | --- | --- | --- | --- |
|  | N | % | N | % | N | % |
| **Full sample** | 3345 | 100 | 1565 | 46.79 | 1780 | 53.21 |
| **Baseline decay (treatable teeth)** | 497 | 14.86<br>(13.7, 16.1) | 221 | 14.12<br>(12.4, 15.8) | 276 | 15.50<br>(13.8, 17.2) |
| **Preexisting sealants** | 1665 | 49.78 | 688 | 43.96 | 977 | 54.89 |
| **Sex (male)** | 1686 | 50.40 | 778 | 49.71 | 908 | 51.01 |
| **Race/Ethnicity** |  |  |  |  |  |  |
| **Hispanic/Latino** | 36 | 1.08 | 13 | 0.83 | 23 | 1.29 |
| **Black** | 30 | 1.97 | 16 | 1.02 | 14 | 0.79 |
| **White** | 868 | 25.95 | 346 | 22.11 | 522 | 29.33 |
| **Asian** | 29 | 0.87 | 10 | 0.64 | 19 | 1.07 |
| **More than one** | 19 | 0.57 | 10 | 0.64 | 9 | 0.51 |
| **Other** | 10 | 0.30 | 1 | 0.06 | 9 | 0.51 |
| **Unreported** | 2353 | 70.34 | 1169 | 74.40 | 1184 | 66.52 |
| **Baseline age (years)** | 7.2 | 1.8 | 7.0 | 1.8 | 7.3 | 1.8 |

**Figure 1:**
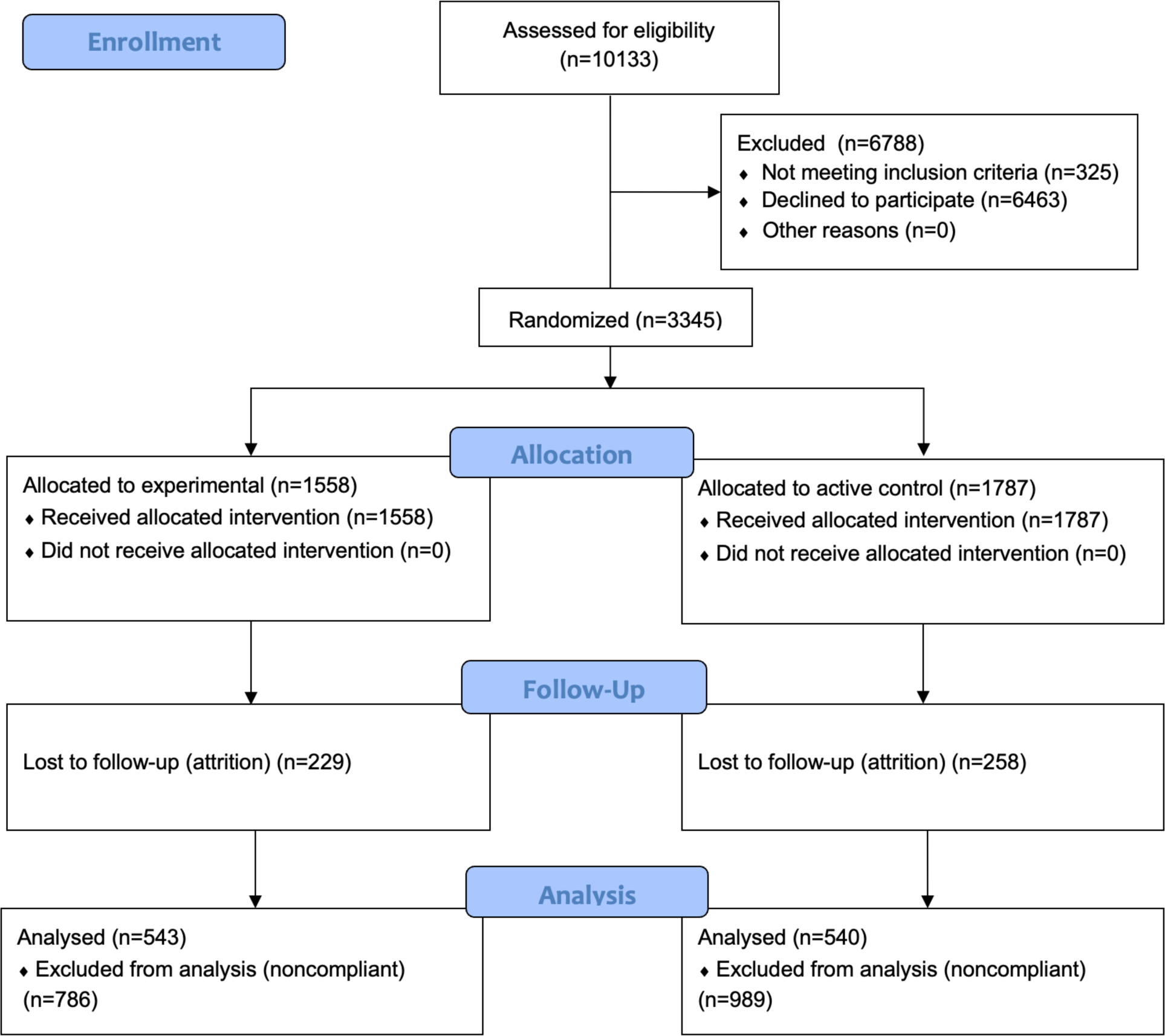
CONSORT Flow Diagram.

There were 487 participants who did not complete a post-baseline observation (229 [47%] in the SDF group, 258 [53%] in the sealant and ART group). There were no significant differences in the prevalence of baseline decay between groups in those lost to follow-up (Supplementary Table 1), however dropouts in the sealant group were more likely to be white, while those in the SDF were more likely to have race/ethnicity unreported. Additionally, 10 (1.8%) participants in the analytic set originally assigned to receive sealants and ART would later crossover to a school assigned to receive SDF, and 22 (3.4%) of those assigned to receive SDF switched to receive sealants and ART. Finally, 184 (28%) participants in the SDF group presented at a follow-up observation with dental sealants (indicative of having received them from an external provider), 52 (13%) of which were observed at the first post-baseline visit. In these cases, proceeding observations for noncompliant participants were removed from analysis.

For the remaining subjects (n=1083; 543 in the experimental group and 540 in the active control), the prevalence of untreated decay was 16.7% (95% CI = 14.5, 18.9), or 14.7% (95% CI = 11.7, 17.7) in the SDF group and 18.7% (95% CI = 15.4, 22.0) in the sealant and ART group. A total of 732 (68%) subjects had a maximum of one post-baseline observation, 138 (13%) had two, 108 (10%) had three, and 66 (6%) had four (data not shown). The average number of days between each of these observational periods was 203, 351, 370, and 321, respectively (3.4 years, data not shown). Less than 4% of study participants had more than four post-baseline observations. The difference in caries prevalence six months after initial treatment was 0.8% (95% CI = -5.4, 3.9), below the non-inferiority threshold. The total post-baseline number of teeth with caries (Table 2) observed in the SDF group was 210 over 186,235 days, whereas tooth-level caries incidence in the sealant and ART group was 179 over 281,154 days, resulting in a crude incidence rate difference of 0.0005 (95% CI = .0003, .0007) and a ratio of 1.77 (95% CI = 1.44, 2.17), indicating higher incidence in the SDF group.

**Table 2:** Caries prevalence and incidence over time (for teeth receiving treatment)

|  | Caries Prevalence |  |  |  | Caries Incidence |  |
| --- | --- | --- | --- | --- | --- | --- |
|  | Sealant |  | SDF |  | Sealant* | SDF* |
| **Obs. # (Exp/Control)** | N | % | N | % | N | N |
| **1 (543/540)** | 101 | 18.70 | 80 | 14.73 | 242 | 183 |
| **2 (543/540)** | 63 | 11.67 | 67 | 12.43 | 131 | 150 |
| **3 (145/202)** | 19 | 9.41 | 24 | 16.55 | 33 | 42 |
| **4 (78/133)** | 4 | 3.01 | 8 | 10.26 | 7 | 10 |
| **5 (29/76)** | 4 | 5.26 | 5 | 17.24 | 6 | 8 |
| **6 (5/34)** | 1 | 2.94 | 0 | 0 | 1 | 0 |

*\*Days of observation for participants in the sealant and ART group was 281,154. Days of observation for participants in the SDF group was 186,235.*

In adjusted analyses, the weighted risk difference (Table 3) for the number of carious teeth between the two groups was not significantly different (B = .0003, 95% CI = -0.0001, 0.0008). For caries prevalence (Table 4), the overall odds significantly decreased with each observational visit (OR = 0.75, 95% CI = 0.64, 0.88). Compared to those receiving sealants and ART, the odds of decay in the SDF group was 1.35 (95% CI = 0.86, 2.11; 90% CI = 0.92, 1.97), below the non-inferiority threshold. Similarly, the incidence of caries decreased over time (IRR = 0.82, 95% CI = 0.75, 0.90), and there were no significant differences between treatment groups (IRR = 1.19, 95% CI = 0.81, 1.74).

**Table 3:**
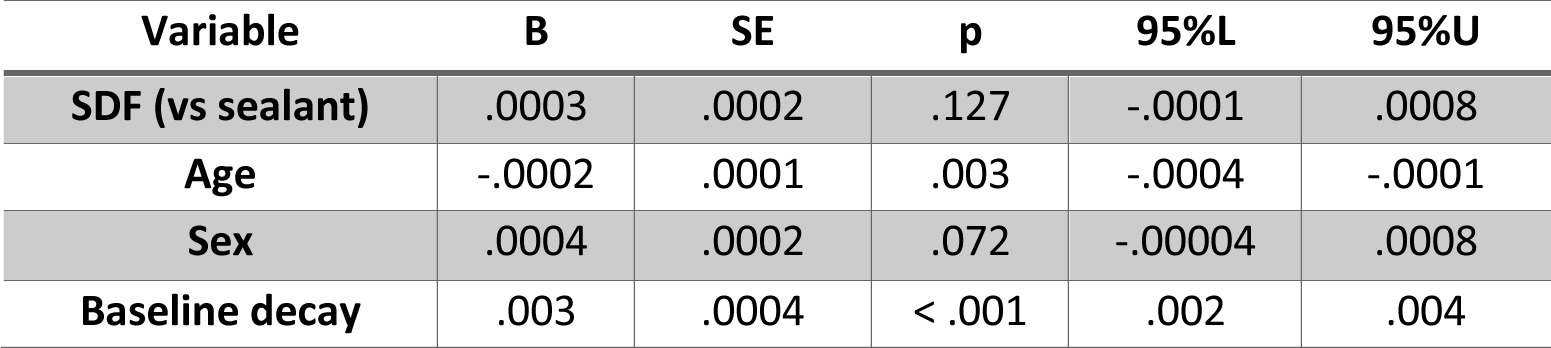
Weighted risk difference of dental caries comparing SDF to sealant participants.

| **Variable** | **B** | **SE** | **p** | **95%L** | **95%U** |
| --- | --- | --- | --- | --- | --- |
| **SDF (vs sealant)** | .0003 | .0002 | .127 | -.0001 | .0008 |
| **Age** | -.0002 | .0001 | .003 | -.0004 | -.0001 |
| **Sex** | .0004 | .0002 | .072 | -.00004 | .0008 |
| **Baseline decay** | .003 | .0004 | < .001 | .002 | .004 |

**Table 4:** Prevalence of untreated caries, adjusted.

| Variable | OR | SE | p | 95%L | 95%U |
| --- | --- | --- | --- | --- | --- |
| **Observational visit** | 0.75 | .06 | < .001 | 0.64 | 0.88 |
| **SDF (vs sealant)** | 1.35 | .31 | .194 | 0.86 | 2.11 |
| **Baseline decay** | 145.0 | 50.1 | < .001 | 78.0 | 288.5 |
| **Number of fillings** | 0.77 | .03 | < .001 | 0.70 | 0.83 |

**Table 5:** Incidence of untreated caries, adjusted.

| Variable | IRR | SE | p | 95%L | 95%U |
| --- | --- | --- | --- | --- | --- |
| **Observational visit** | 0.82 | .04 | < .001 | 0.75 | 0.90 |
| **SDF (vs sealant)** | 1.19 | .23 | .373 | 0.81 | 1.74 |
| **Baseline decay** | 50.25 | 8.93 | < .001 | 35.48 | 71.18 |
| **Number of fillings** | 0.80 | .02 | < .001 | 0.76 | 0.85 |

In ITT analyses (Supplementary Table 2), the odds of decay significantly decreased with each observational visit (OR = 0.70, 95% CI = 0.61, 0.80). The odds comparing SDF to sealants and ART was slightly attenuated compared to per-protocol analyses, with an odds ratio of 1.28 (95% CI = 0.97, 1.70; 90% CI = 1.02, 1.62). Caries prevalence between treatment groups remained nonsignificant and non-inferior.

Results from multiplicative models in adjusted analyses indicate that those in the SDF group had a significantly higher rate of caries over time compared to those receiving sealants (IRR = 1.27, 95% CI = 1.06, 1.53). Those with baseline decay had lower rates over time (IRR = 0.45, 95% CI = 0.38, 0.54), reflecting the effect of the initial treatment of existing lesions.

## Discussion

In this randomized clinical trial and school-based caries prevention program, we observed no differences in caries prevalence six months after initial treatment with either silver diamine fluoride or dental sealants with atraumatic restorations. In adjusted analyses, the overall prevalence of untreated decay and the incidence rate of new caries significantly decreased with each observational visit, and SDF was non-inferior to dental sealants and ART over 3.5 years. Results for adjusted ITT analyses were similar to per-protocol. We conclude that integrating silver diamine fluoride into school-based caries prevention may be an effective strategy to increase access to care and mitigate the burden of caries. However, due to many study disruptions from COVID-19, observed effects may be unreliable beyond short-term findings.

Growing evidence supports the suitability of silver diamine fluoride for the primary prevention of dental caries. Prior estimates of the incidence of caries in children treated with SDF ranges between 23% to 52% less than those receiving placebo ^24^, compared to 65% to 70% less in those receiving dental sealants ^24^. Similarly, a 2023 randomized trial concluded that SDF was more effective in preventing caries in primary teeth compared to fluoride varnish ^31^, a review of SDF prevention studies estimated a preventive fraction of 61% for SDF compared to controls and a tooth number needed to treat of 4 and 12.1 for preventing caries in the primary dentition and first permanent molars, respectively ^24^, and no differences were found between SDF, fluoride varnish, and resin sealants in the two-year incidence of caries in permanent molars ^32^. More recently, a large pragmatic trial conducted in predominantly low-income minority children living in urban areas reported nearly identical prevalence and incidence of caries over time in participants receiving SDF or dental sealants ^25,33^. Caries diagnosis in this prior study was determined by ICDAS scores of >4. In contrast, the present results expand caries diagnosis to ICDAS scores of >3, including lesions with underlying dark shadow from dentin with or without enamel breakdown. This additional criteria may have contributed to the higher estimates of caries prevalence and incidence in the presented study.

School-based caries prevention with dental sealants reduces caries risk and disability-adjusted life years, and are cost-effective ^34^. However, this approach can be underused due to prohibitive costs or local policies requiring that dentists be present in a supervisory capacity ^18^. Considering the other benefits of SDF including cost-effectiveness ^21^, patient tolerability ^35^, efficiency of treatment time ^22^, and relative ease of application ^24^, integrating silver diamine fluoride into school-based programs may result in more children being treated in less time and cost. Indeed, prior studies note particular utility for SDF in dental public health programs and community approaches to caries prevention, especially amongst those in dental shortage areas or where sealants are otherwise infeasible ^32^. Additionally, while the US Preventive Services Task Force currently holds that there is insufficient data to assess the benefits and harms of oral disease prevention conducted by primary care clinicians ^36^, there is preliminary evidence that SDF can be effectively applied by physicians ^37^ and nurses ^23,25^.

Prior data indicates that the global distribution of dentists is inequitable, with creative workforce development being a critical need to achieve universal health coverage ^38^. The simplicity of SDF application can lead to efficient increases in the oral health workforce. For example, with training and support, schools in extremely under-resourced areas may be able to leverage existing school nurse personnel and SDF in creating a sustainable approach to school-based caries prevention. Alternatively, the use of SDF in school-based caries prevention can be implemented as an early interventional strategy, providing efficient options for caries prevention and control while preserving more limited and comprehensive resources, such as dental sealants, for children with only permanent dentition.

There are multiple limitations in this study that limit the validity and generalizability of our results. First, our outcomes for decay did not consider prior states of individual teeth, and as a result could reflect decay emerging either from previously sound teeth (primary prevention failure) or those that were decayed and treated (secondary prevention or caries management failure). It is also possible that permanent teeth erupting between observational visits could have developed decay prior to treatment. Additionally, disruptions in study activities due to school closures resulting from COVID-19 meant that the original protocol for treatment frequency could not be followed, and the planned biannual treatment was inconsistent. As a result, the time between observations ranged from 7 to 13 months. These disruptions also negatively influenced follow-up in study participants: the overall follow-up rate was approximately 33%, there were considerable reductions in the number of participants who were observed at successive visits, and a number of schools were unable to be seen for follow-up observations. There was also a high prevalence of preexisting sealants in both groups, and their exclusion substantially reduced the number of participants viable for analysis. Although the crossover rate between treatment groups was minimal, a large proportion of children in the SDF group were noncompliant due to receiving dental sealants from providers outside of the school-based program. This exclusion resulted in greater loss to follow-up in the experimental group, and could further bias results. As noncompliance primarily resulted in SDF participants receiving the interventions provided in the active control arm via an external provider, it might be expected that the effect of treatment assignment is more biased towards the null than the effect of treatment itself, and this was indeed observed. While our methods considered a number of approaches to mitigate these limitations, it is likely that unobservable characteristics introduced bias into our results, and the presented findings must be considered with caution.

In conclusion, despite negative impacts of COVID-19, there may be potential benefits of incorporating silver diamine fluoride into school-based caries prevention in order to increase access to critical oral healthcare for high-risk, low-resource populations. Further study is warranted.

## Data Availability

Data available: Yes
Data types: Data dictionary
How to access data: Data dictionaries will be available to interested researchers upon request to the authors
When available: beginning date: 06-01-2025
Supporting Documents
Document types: Informed consent form
How to access documents: Informed consent forms will be available to interested researchers upon request to the authors
When available: beginning date: 06-01-2025
Additional Information
Who can access the data: Interested researchers upon request to the authors
Types of analyses: For any purpose
Mechanisms of data availability: After approval of a proposal and a signed data access agreement.

## Author Contributors

*Concept and design:* RRR, RN

*Acquisition, analysis, or interpretation of the data:* RRR, RN

*Drafting of the Manuscript*: RRR

*Critical review of manuscript*: RRR, RN

*Statistical Analysis*: RRR

*Obtained funding*: RRR, RN

*Administrative, technical, or material support*: RRR, RN

*Supervision:* RRR, RN

## Data Sharing

**Data available:** Yes

**Data types:** Data dictionary

**How to access data:** Data dictionaries will be available to interested researchers upon request to the authors

**When available:** beginning date: 06-01-2025

## Supporting Documents

**Document types:** Informed consent form

**How to access documents:** Informed consent forms will be available to interested researchers upon request to the authors

**When available:** beginning date: 06-01-2025

## Additional Information

**Who can access the data:** Interested researchers upon request to the authors

**Types of analyses:** For any purpose

**Mechanisms of data availability:** After approval of a proposal and a signed data access agreement.

## Acknowledgements

TBA

---

**Supplementary Table 1:** Characteristics of participants lost to follow-up.

|  | Overall |  | SDF |  | Sealants |  |
| --- | --- | --- | --- | --- | --- | --- |
|  | N | % | N | % | N | % |
| **Full sample** | 487 | 100 | 229 | 47.02 | 258 | 52.98 |
| **Baseline decay (treatable teeth)** | 94 | 19.30<br>(15.8, 22.8) | 42 | 18.34<br>(13.3, 23.4) | 52 | 20.16<br>(15.2, 25.1) |
| **Preexisting sealants** | 1716 | 46.76 | 703 | 41.62 | 1013 | 51.14 |
| **Sex (male)** | 259 | 53.18 | 130 | 56.77 | 129 | 50.0 |
| **Race/Ethnicity** |  |  |  |  |  |  |
| **Hispanic/Latino** | 10 | 2.05 | 4 | 1.75 | 6 | 2.33 |
| **Black** | 11 | 2.26 | 6 | 2.62 | 5 | 1.94 |
| **White** | 310 | 63.66 | 125 | 54.59 | 185 | 71.71 |
| **Asian** | 13 | 2.67 | 4 | 1.75 | 9 | 3.49 |
| **More than one** | 5 | 1.03 | 3 | 1.31 | 2 | 0.78 |
| **Other** | 2 | 0.41 | 0 | 0 | 2 | 0.78 |
| **Unreported** | 136 | 27.93 | 87 | 37.99 | 49 | 18.99 |
| **Baseline age (years)** | 6.6 | 1.72 (SD) | 6.7 | 1.73 (SD) | 6.5 | 1.71 (SD) |

**Supplementary Table 2:** Prevalence of dental caries, adjusted (ITT)

| Variable | OR | SE | p | 95%L | 95%U |
| --- | --- | --- | --- | --- | --- |
| **Observational visit** | 0.70 | .05 | < .001 | 0.61 | 0.80 |
| **SDF (vs sealant)** | 1.28 | .18 | .079 | 0.97 | 1.70 |
| **Baseline decay** | 54.13 | 10.08 | < .001 | 37.58 | 77.97 |
| **Number of fillings** | 0.83 | .03 | < .001 | 0.78 | 0.89 |

